# Characteristics, clinical outcomes, and mortality of older adults living with HIV receiving antiretroviral treatment in the sub-urban and rural areas of northern Thailand

**DOI:** 10.1101/2022.06.27.22276956

**Authors:** Linda Aurpibul, Patumrat Sripan, Wason Paklak, Arunrat Tangmunkongvorakul, Amaraporn Rerkasem, Kittipan Rerkasem, Kriengkrai Srithanaviboonchai

## Abstract

Since the introduction of antiretroviral treatment (ART), people living with HIV worldwide live into older age. This study described the characteristics, clinical outcomes, and mortality of older adults living with HIV (OALHIV) receiving ART from the National AIDS program in northern Thailand. Participants aged ≥ 50 years were recruited from the HIV clinics in 12 community hospitals. Data were obtained from medical records and face-to-face interviews. In 2015, 362 OALHIV were enrolled; their median (interquartile range) age and ART duration were 57 years (54-61), and 8.8 years (6.4-11.2), respectively. At study entry, 174 (48.1%) had CD4 counts ≥ 500 cells/mm^3^; 357 of 358 (99.6%) with available HIV RNA results were virologic-suppressed. At the year 5 follow-up, 39 died, 11 were transferred to other hospitals, 3 were lost to follow-up, and 40 did not contribute data for this analysis, but remained in care. Among the 269 who appeared, 149 (55%) had CD4 counts ≥ 500 cells/mm^3^, and 227/229 tested (99%) were virologic-suppressed. The probability of 5-year overall survival (OS) was 89.2% (95% confidence interval, CI 85.4-92.1%). A significantly low 5-year OS (66%) was observed in OALHIV with CD4 counts < 200 cells/mm^3^ at study entry. The most common cause of death was organ failures in 11 (28%), followed by malignancies in 8 (21%), infections in 5 (13%), mental health-related conditions in 2 (5%), and unknown in 13 (33%). In OALHIV with stable HIV outcomes, monitoring of organ functions, cancer surveillance, and mental health screening are warranted.

## Introduction

With effective antiretroviral treatment (ART), people living with HIV worldwide survive and stay healthy. Data from developed countries reported a substantial reduction in the mortality rate of people living with HIV (PLHIV) and an increasing life expectancy approaching that of the general population (1, 2). In Thailand, the universal coverage National AIDS Program provides HIV care and ART free of charge since 2007 (3). The thresholds set for starting treatment have been updated since then, from CD4 < 200 cells/mm^3^ during 2008-2010, CD4 < 350 cells/mm^3^ from 2011-2013, and any CD4 level since 2014 (4). The practice following this guideline led to an overall increase in the number of PLHIV accessing ART; in 2019, 80% of adults living in Thailand were on received treatment (5). This will create a demographic shift in PLHIV care, as everyone could live longer.

In 2019, the HIV incidence in Thailand was 0.15 per 1000 population; with 470,000 adults aged 15 years and older living with HIV, and the estimated number of deaths due to AIDS ranged between 9,700 to 15,000 per year (5). Similar to other parts of the world, since early in the epidemic, the proportion of people older than age 50 years so-called older adults living with HIV (OALHIV) in Thailand increased with time. OALHIV were more likely to be diagnosed late due to perception of low HIV-acquisition risk, limited knowledge about the disease (6), misunderstandings about HIV/AIDS, as well as stigmatization, and discrimination in the past (7). The cohort data from Public Health England mentioned late diagnosis as a strong predictor of death (1). Moreover in the past decade, PLHIV needed to wait until immunosuppressed to be eligible for ART (4). Late ART initiation at low baseline CD4 counts is associated with a slow and low rate of CD4 recovery after the commencement of ART (8, 9).

The analysis of the Thai National AIDS Program data from 2008 to 2014 revealed a decrease in overall life expectancy after ART initiation in those with older age and lower baseline CD4; however, the median age of the population in that report was only 37 years (3). The studies from South Africa and Uganda documented that despite ART, a higher mortality rate was observed among those older than 50 years when compared to younger age and non-HIV age-matched people, respectively (10, 11). As known that aging is accompanied by physiological changes, and higher risk of non-communicable diseases. Comorbidities including cardiovascular disease, heart/vascular, and malignancy were found to be a major cause of death that were strongly associated with age in the ART collaboration cohort (12). Immune dysfunction and chronic inflammation were associated with increased risk of malignancies (13), while the use of certain ART regimens was associated with cardiovascular risk (14). In the US veterans cohort, the prevalence of medical comorbidities was higher in those with HIV infection, especially in older age, and increase HIV disease severity (15). Frailty, defined as a vulnerability to adverse outcomes in the face of stressors was reported to be more prevalent and occurred earlier in PLHIV compared to populations without HIV, and was associated with adverse clinical outcomes and mortality.(16)

Currently, data on OALHIV in Asia remains scarce. We hypothesize that the HIV treatment outcome and mortality of OALHIV might be different from PLHIV at younger age. This study aimed to describe the characteristics, clinical outcomes, and mortality of OALHIV in the rural and suburban areas in northern Thailand. The study results would enable us to gain insights into the long-term impact of the national HIV program as well as to guide the future management plan on monitoring and screening for conditions that could compromise the HIV treatment outcome in an aging population.

## Materials and methods

### Study population

The prospective cohort study was started in August 2015. The study participants were OALHIV who attended HIV clinics for antiretroviral treatment at 12 community hospitals in Chiang Mai, Thailand. The primary aim of the cohort is to follow the quality of life and health outcomes of OALHIV receiving the national standard of HIV care. At that time potential participants were approached and invited to join the study during routine clinical visits on a first come first serve basis. The inclusion criteria were 1) having a diagnosis of HIV infection, 2) aged ≥ 50 years, and 3) receiving antiretroviral treatment at study enrollment. For each hospital, the oldest patients were approached first. After enrollment, all remained followed in HIV clinics for regular care at 2- or 3-month intervals under the Thai national AIDS program coverage.

### Data collection and tools

Baseline demographic data included age, sex, years of formal education, employment status, family status, monthly income, history of smoking, and alcohol use were collected in 2015 which was the first year of study enrollment. Bodyweight and height were measured at entry and the body mass index (BMI) was calculated. The clinical information was retrieved from medical records by HIV clinic staff at each hospital including duration of treatment, the current regimen used, the most recent CD4 counts, and HIV-RNA level within 12 months before the study entry. The second-round survey was conducted in 2016. The most recent data collection was completed in September-November 2020. During each survey, we reconsented each study participant for their willingness to join. The medical record review for CD4 counts and HIV RNA level. The participant interview session was conducted with each participant to update their demographic characteristics. For participants who did not show up at clinic, their vital status was recorded as dead, or alive but lost-to-follow-up, or alive with transferred out to other hospitals. Data from those who remained in care but were unavailable to come on the day of study follow-up or did not give consent for continuous participation was not retrieved or included in the study analysis.

For those who died in the hospitals, the cause of death and/or final diagnosis before death were reviewed from medical records by HIV clinic staff. For participants who died outside the healthcare facilities, medical record was also reviewed for the most recent diagnosis which might be relevant to their mortality. In addition, their next-to-kin relatives were contacted by HIV clinic staff who were familiar with their families. They were asked for coming to clinic for informed consent for an interview. The verbal autopsy questionnaire was used as a guide (17). Data from the last clinic visit, hospital records, and laboratory results were reviewed to gather as much information as possible to identify the cause of death for each participant. The primary causes of death were finally determined by two investigators who are medical doctors (KS and LA). In cases with insufficient data, the primary cause of death was marked as unknown.

### Ethics statement

The study was approved by the institutional review board at Research Institute for Health Sciences, Chiang Mai University (Certificate approval number 35/2020). Written informed consent was obtained from each participant before enrollment.

### Statistical analysis

Baseline demographic characteristics of study participants were reported as medians and interquartile ranges (IQRs) for continuous variables, or numbers and percentages for categorical variables. The comparison was made between those with CD4 counts < 200, 200-499, and ≥ 500 cells/mm^3^ using Kruskal-Wallis test for continuous variables and Fisher’s exact test for categorical variables.

CD4 counts at the current 5th-year follow-up were categorized to 4 groups:, <200, 200-349, 350-499 and ≥500 cells/ mm^3^. The proportions of male, lifetime smoking, and alcohol use in the past year in group of CD4 were compared using Fisher’s exact test. The Kruskal–Wallis Test was used to compare median age, years of formal education, duration after HIV diagnosis, duration on ART, and CD4 counts at study enrollment between the group of CD4 at the 5^th^ year.

The survival rates were estimated using the Kaplan-Meier method. Overall 5-year survival was defined as the study entry to the date of death from any causes. The probabilities of 5-year overall survival (OS) were calculated for each CD4 group and compared using Log rank test. Factors associated with the probability of 5-year OS were determined and reported as hazard ratio (HR). Factors with significantly associated indicated by p< 0.05 or with clinically relevant were included in the adjusted model. The adjusted hazard ratio (aHR) was reported. The statistical significance was defined at p<0.05, and all P-values reported in this article are two-sided values, determined using Stata version 14 (StataCorp LP, College Station, TX, USA).

## Results

### Cohort description

In 2015, a total of 364 OALHIV were screened; two were excluded as they have not yet initiated on ART. Three hundred and sixty-two OALHIV were enrolled into the cohort; 155 (43%) were male. Their median age was 57 years (interquartile range, IQR 54-61). There were 262 (72.4%) who had primary school education, while 55 (15.2%) attended secondary school or higher, 45 (12.4%) have not attended school. At the time of study enrollment, 175 (48.3%) were unemployed or have retired, while 74 (20.4%) remained laborers/ employed for wages, 48 (13.3%) worked in agricultural, 12 (3.3%) in government sectors, and 53 (14.6%) had independent jobs. A hundred and thirty-four (37%) reported smoking in their lifetime, and 68 (18.8%) used alcohol in the past year. Seventy-nine (21.8%), 243 (67.1%), and 40 (11.1%) had BMI in low, normal, and overweight range, respectively.

Their median duration on ART was 8.8 years (IQR 6.4-11.2). The most common ART regimens used were non-nucleoside reverse transcriptase inhibitor (NNRTI)-based in 336 (92.8%), others 26 (7.2%) were on the protease-inhibitor (PI)-based regimens. At this study enrollment, 163 (45.0%), and 174 (48.1%) had CD4 counts 200-499, and > 500 cells/mm^3^, respectively; 357 of 358 (99.6%) with available HIV RNA results had virologic suppression within 12 months prior to the study enrollment. Comparison of characteristics between the group with CD4 counts < 200, 200-499, and ≥ 500 cells/mm^3^ are shown in Table 1.

**Table 1.**
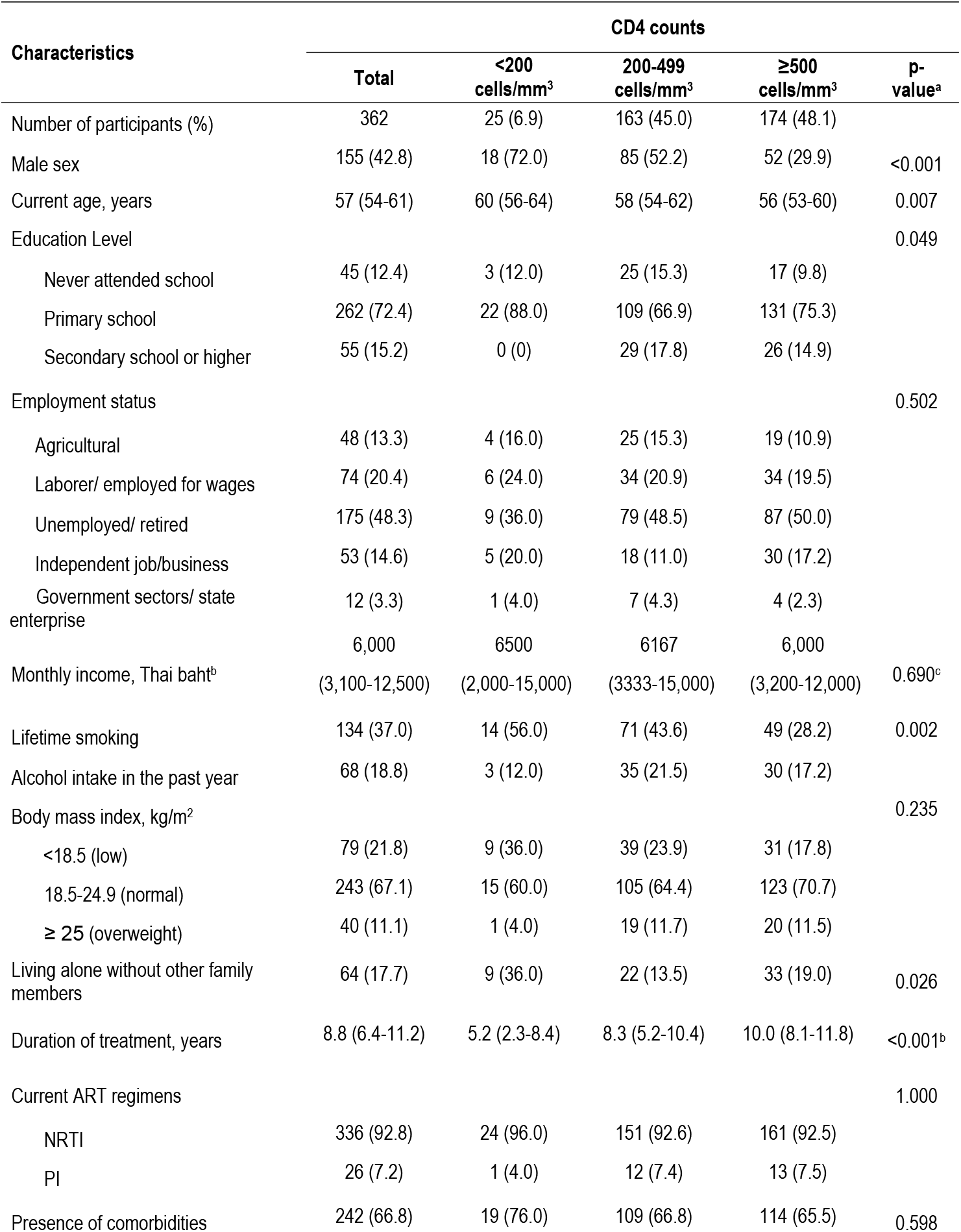
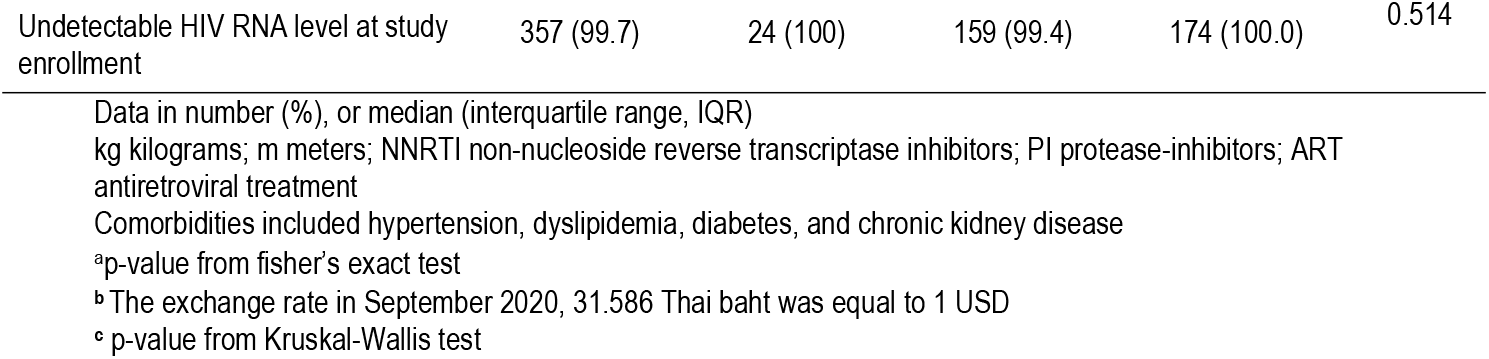
Demographic characteristics of older adults living with HIV stratified by CD4 counts at study enrollment in 2015.

In 2016, there were 325 participants who attended the second-round survey. Among the 37 patients who did not show up; 17 died, 4 were transferred to other hospitals, and 16 remained in care but were unavailable for the survey. In the most recent survey in 2020, there were 269 OALHIV who showed up; 22 died, 7 were transferred to other hospitals, 22 remained in care but were unavailable for the survey, 2 were bedridden (one post-operative and another post-accident), and 3 were lost-to-follow-up. (Fig 1)

**Figure 1.**
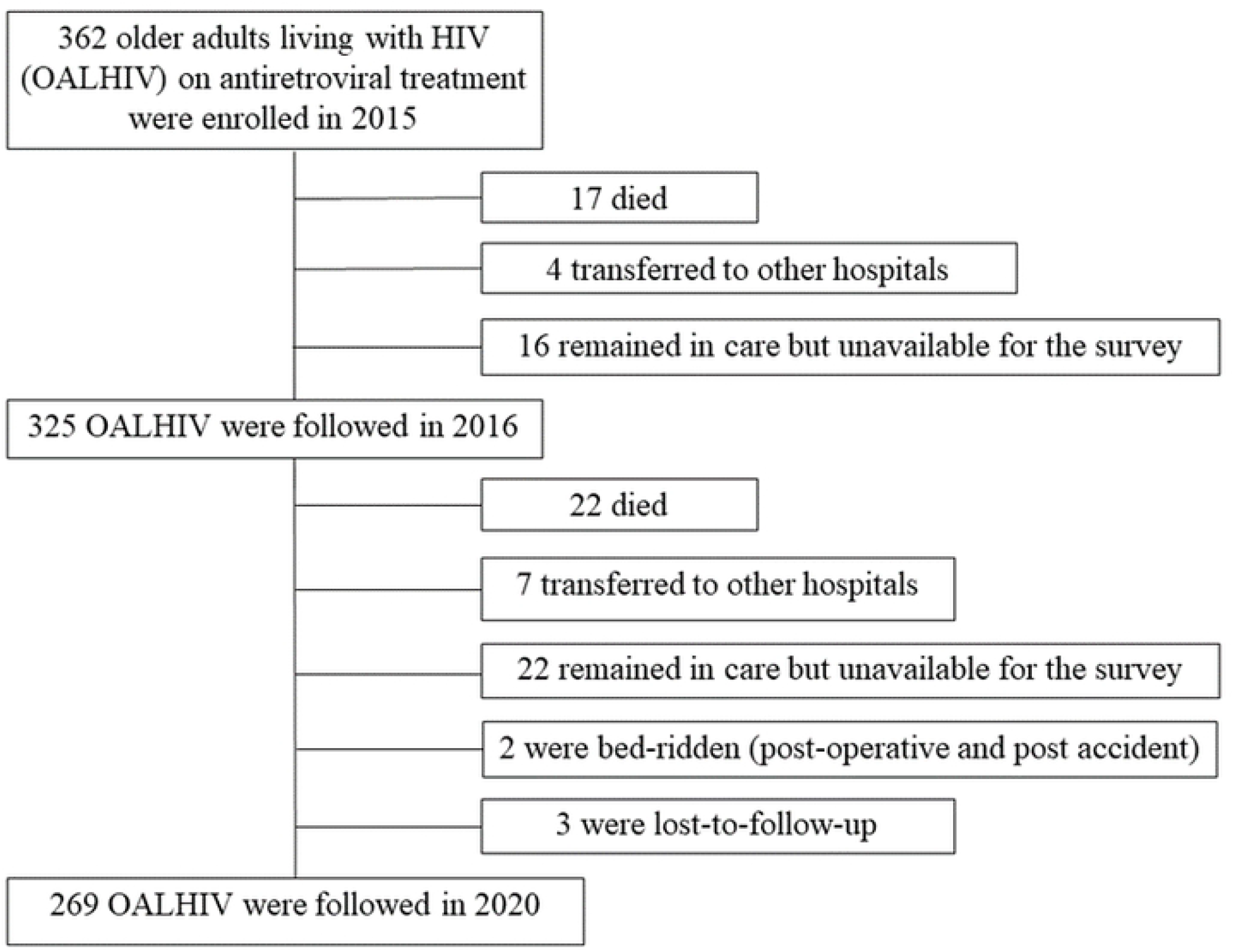
Flow chart of study participants’ status after the study enrollment

### Characteristics of study participants in the most recent survey

Of the 269 participants, 110 (41%) were male. Their current median age was 61 years (IQR 59-65). There were 149 (55%), 61 (23%), 47 (17%), and 12 (5%) with CD4 counts ≥ 500, 350-499, and < 200 cells/mm^3^, respectively. The group with CD4 counts < 200 mm^3^ were at older age (median 64 years), 11 out of 12 (83%) were male, 70% were lifetime smokers, and had low CD4 counts (median 222 cells/mm^3^) at the study entry. Comparison of their characteristics are shown in Table 2.

**Table 2.**
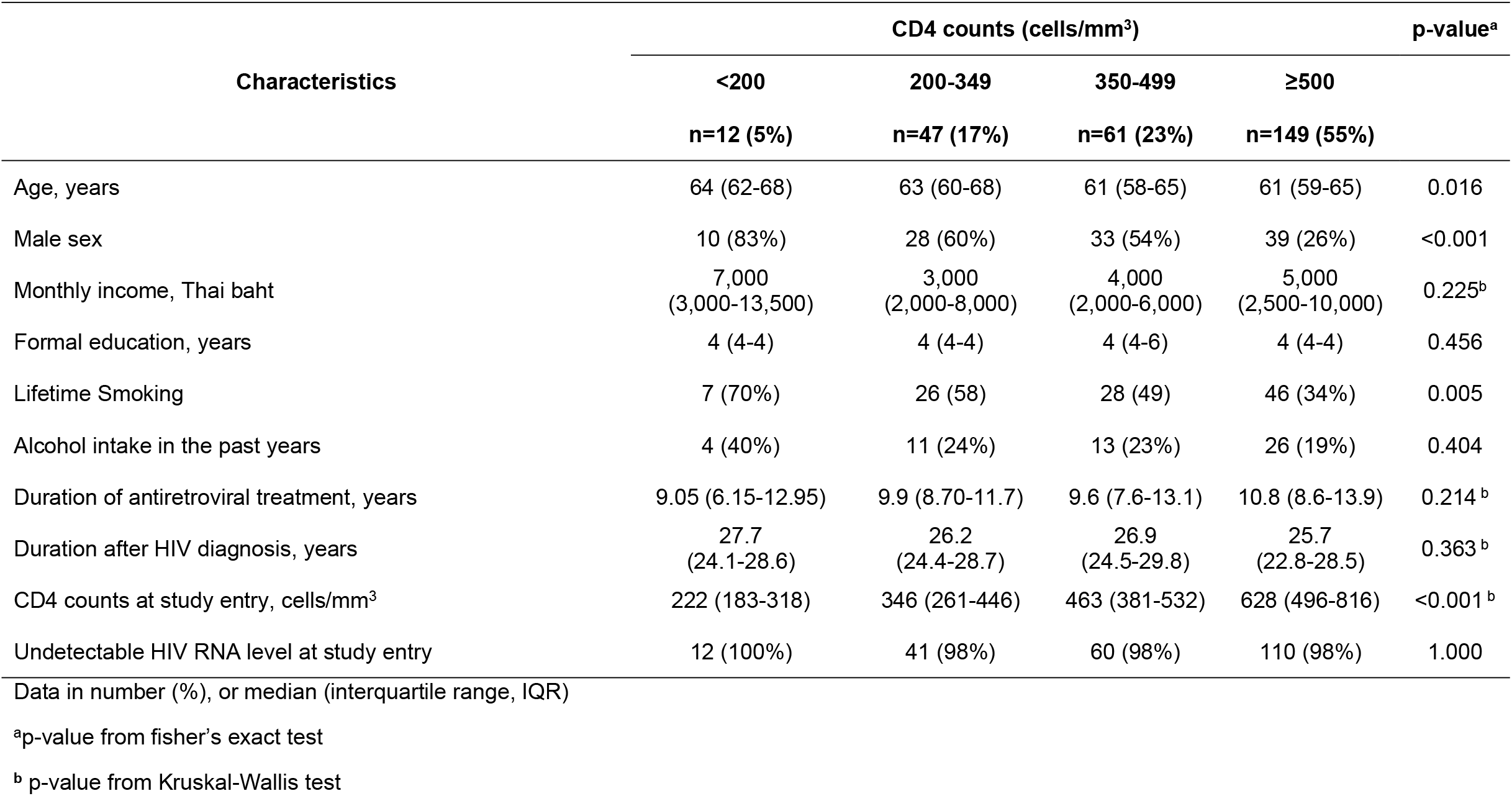
CD4 distribution and associated factors in older adults living with HIV at 5-year follow-up in 2020 (n=269)

### CD4 and HIV RNA trends

The CD4 distributions of OALHIV from 2017-2020 stratified by sex was depicted in Fig 2. During the current survey in 2020, the median CD4 counts was 484 cells/ mm^3^ (IQR 339-634); with slightly more than half (55%) of participants had CD4 cell count ≥ 500 cells/mm^3^. Female had a significantly higher median CD4 counts when compared to male OALHIV both study entry (556 vs. 438 cells/ mm^3^, respectively; p < 0.001) and at year 5 follow-up (539 vs. 416 cells/ mm^3^, respectively; p < 0.001). The HIV RNA level of OALHIV was illustrated in Fig 3. More than 90% had virologic suppression since the study entry to year 5 follow-up with no significant difference between sex.

**Figure 2.**
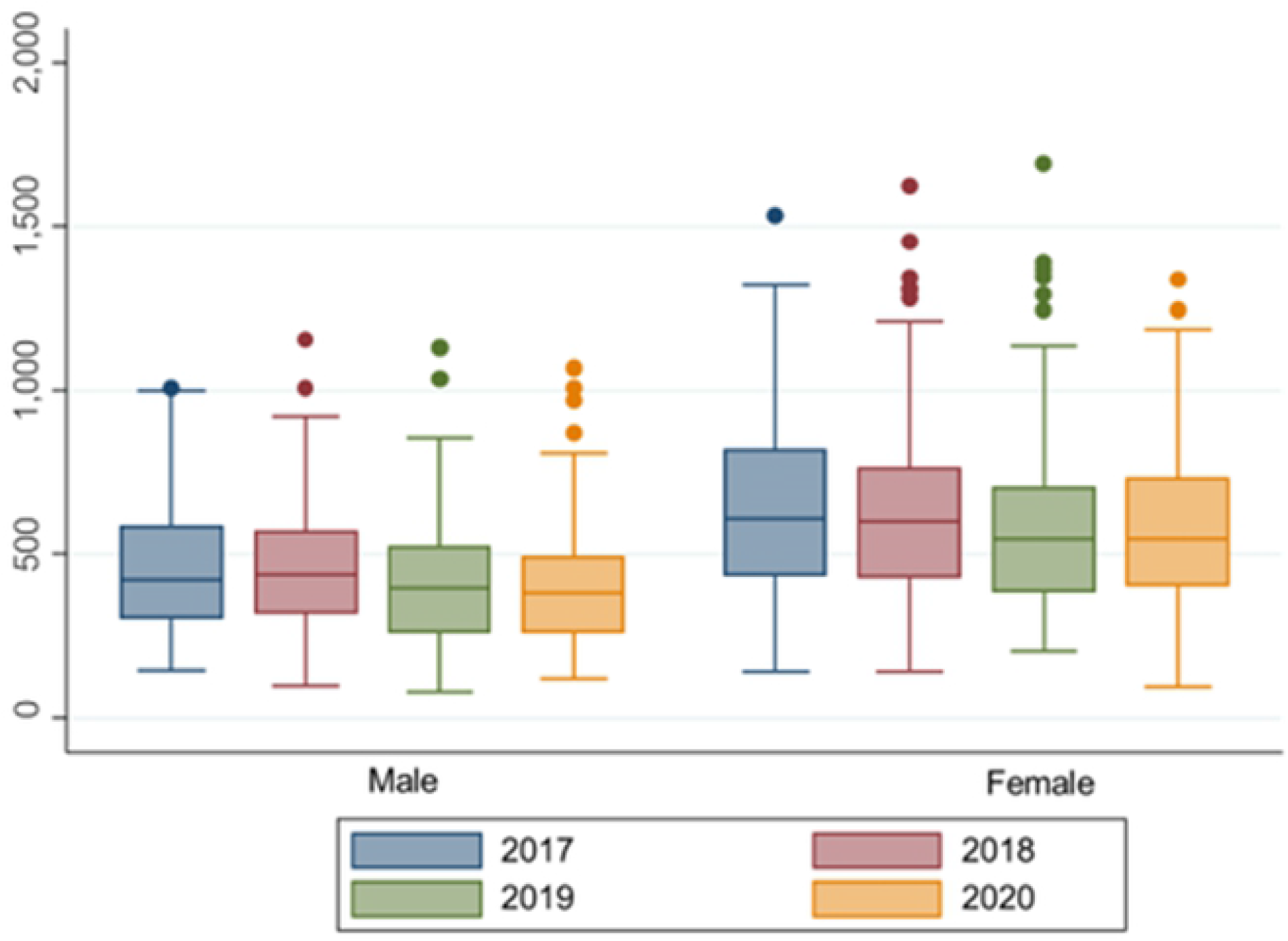
CD4 distributions of older adults living with HIV in 2017-2020 stratified by sex

**Figure 3.**
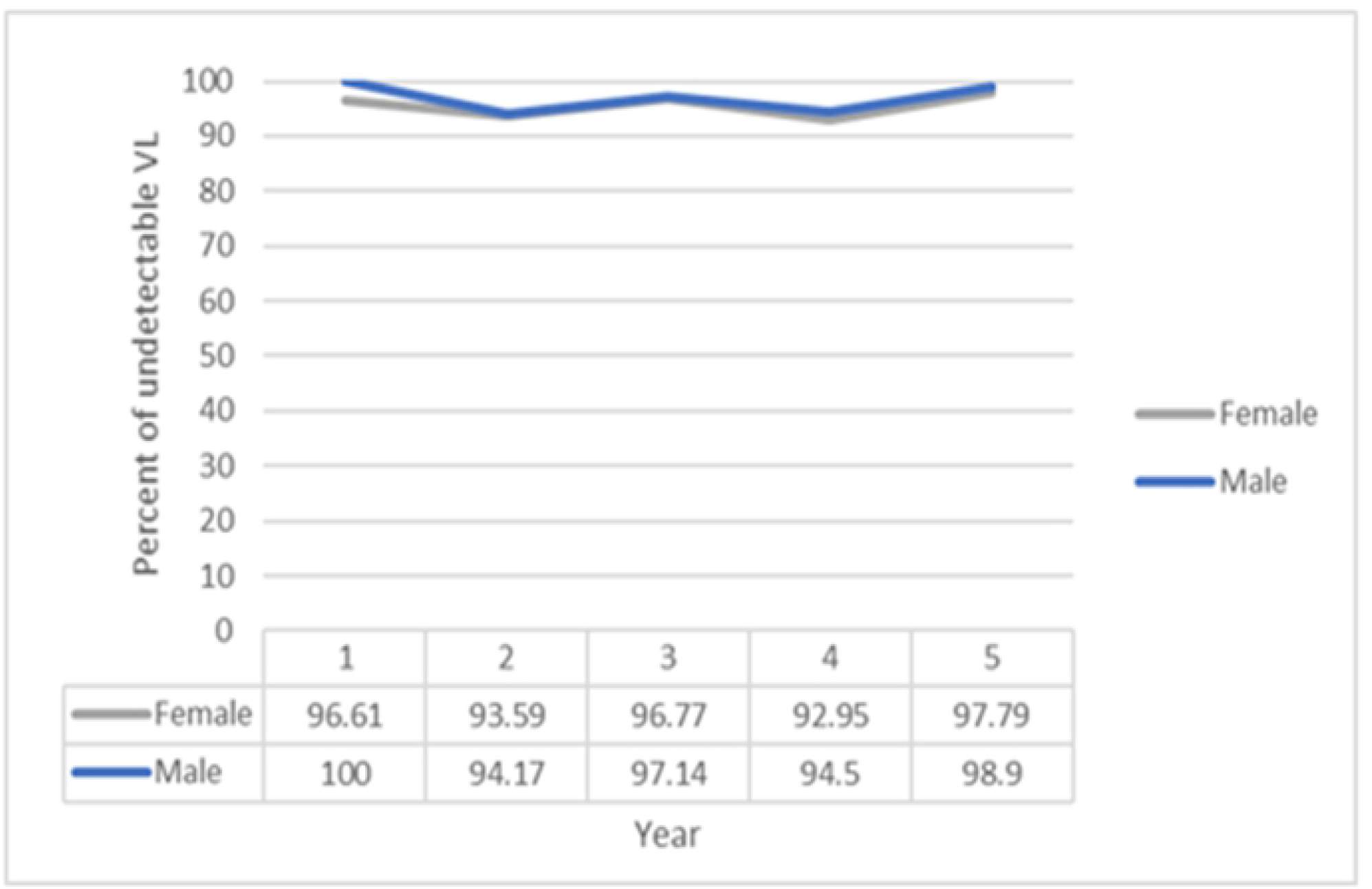
Percentage of undetectable HIV RNA in older adults living with HIV on antiretroviral treatment from 2016-2020 stratified by sex

### Overall survival and Mortality

In the 5-year follow-up time after study entry, a total 39 participants died. The probability of 5-year OS was 89.2% (95% confidence interval, CI 85.4-92.1%). The Kaplan-Meier survival estimate of all participants is shown in Fig 4. The Log-rank test revealed that OALHIV with CD4 counts < 200 cells/mm^3^ at study entry had a significant lower probability of 5-year OS (66.0%) when compared to other two groups with CD4 counts 200-499 cells/mm^3^ (89.4%), and ≥ 500 cells/mm^3^ (92.5%); p< 0.001 (Table 3). The probabilities of 5-year OS in each CD4 group are shown in Fig 5.

**Table 3.** Predictors of mortality in 5-year follow-up time among older adults living with HIV.

| Characteristics | HR | 95% CI | p-value | aHR | 95% CI | p-value |
| --- | --- | --- | --- | --- | --- | --- |
| CD4 counts at study entry |  |  |  |  |  |  |
| <200 cells/mm<sup>3</sup> | 5.45 | 2.23-13.35 | <0.001 | 4.12 | 1.56-10.85 | 0.004 |
| 200-500 cells/mm<sup>3</sup> | 1.45 | 0.69- 3.07 | 0.328 | 1.37 | 0.64-2.96 | 0.419 |
| >500 cells/mm<sup>3</sup> (ref) | 1 |  |  | 1 |  |  |
| Age | 1.06 | 1.01-1.12 | 0.011 | 1.04 | 0.99-1.10 | 0.104 |
| Presence of Comorbidities | 1.04 | 0.52-2.07 | 0.918 | 0.08 | 0.39-1.63 | 0.535 |
| Lifetime Smoking | 1.14 | 0.58-2.22 | 0.707 | 0.90 | 0.45-1.80 | 0.770 |
| Body mass index |  |  |  |  |  |  |
| 18.5-24.9 kg/m<sup>2</sup> (ref) | 1 |  |  |  |  |  |
| <18.5 kg/m<sup>2</sup> | 1.82 | 0.89-3.69 | 0.099 | 1.38 | 0.65-2.91 | 0.405 |
| ≥ 25 kg/m<sup>2</sup> | 0.89 | 0.27-2.97 | 0.852 | 0.99 | 0.29-3.33 | 0.982 |
| Living alone without other family members | 2.01 | 0.99-4.09 | 0.053 | 1.50 | 0.70-3.22 | 0.297 |

HR hazard ratio; aHR adjusted hazard ratio; CI confidence interval

**Figure 4.**
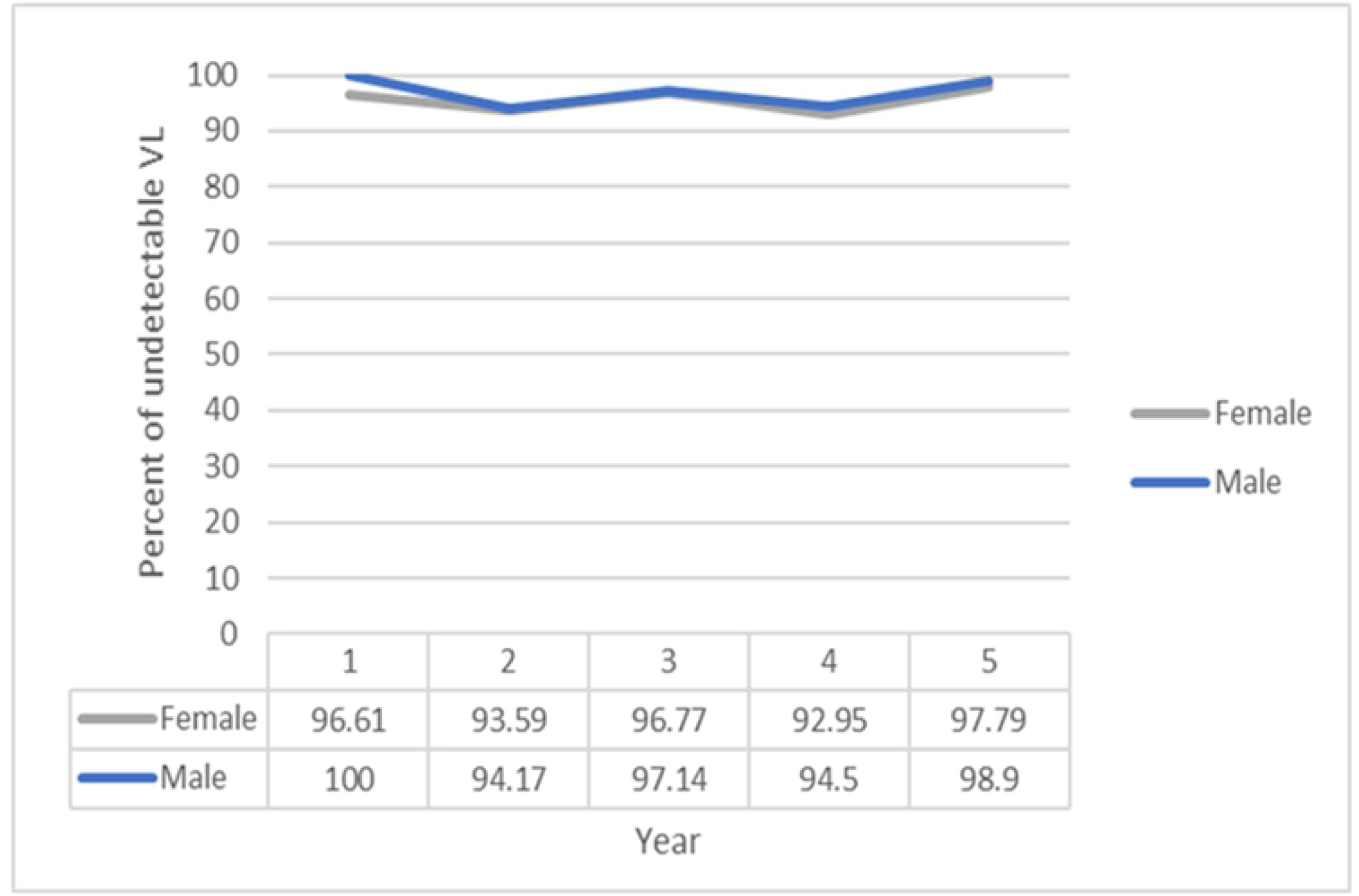
Percentage of undetectable HIV RNA in older adults living with HIV on antiretroviral treatment from 2016-2020 stratified by sex

**Figure 5.**
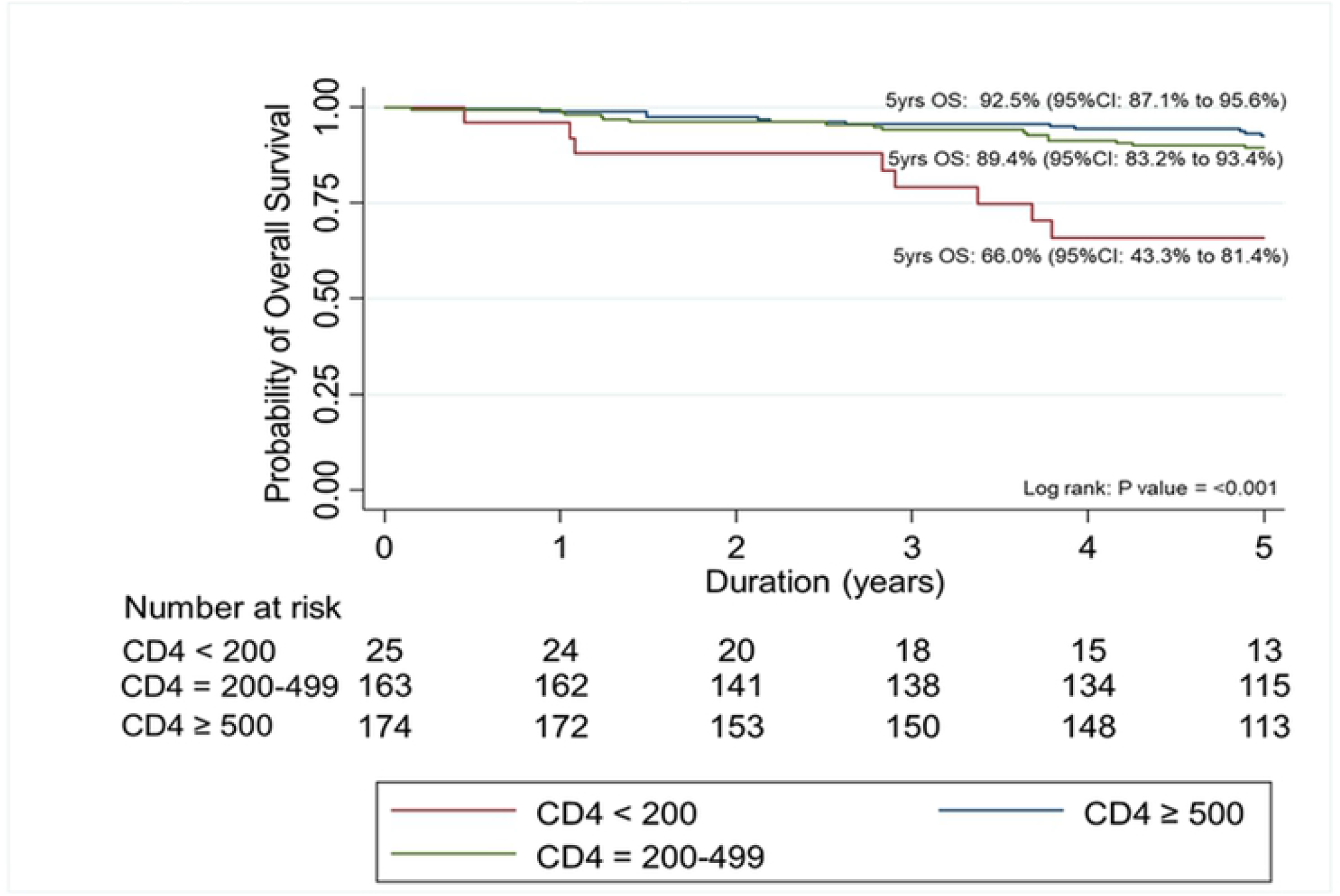
Kaplan-Meier estimates the probability of 5-year overall survival for older adults living with HIV on antiretroviral treatment by CD4 counts at study entry

Of those who died, 17/39 (43.5%) were female. Their median age at death was 62 years (IQR 58-67); 14 (36%), 19 (49%), and 6 (15%) were < 60, 60-69, and 70-79 years at the time of death, respectively. Their most recent CD4 counts were < 200 cells/mm^3^ in 8 (20.5%), and all were virologic-suppressed at the last measure prior to death. The causes of death were obtained from medical records in 11 (28%), from incomplete medical records and interview with relatives in 10 (26%), from interview with relatives only in 6 (15%), and not available in 12 (31%). The most common cause of death was organ failures in 11 (28%), followed by malignancies in 8 (21%), infections in 5 (13%), mental health related conditions in 2 (5%), and unknown in 13 (33%). The causes of death in each age group is shown in Table 4.

**Table 4.** Causes of death in older adults living with HIV stratified by age at death (n=39)

| Causes of death | 50-59 years | 60-69 years | 70-79 years | Total |
| --- | --- | --- | --- | --- |
| Organ failures |  |  |  | 11 |
| Renal | 3 | 2 |  |  |
| Liver |  | 2 |  |  |
| Heart | 1 | 1 |  |  |
| Respiratory |  | 2 |  |  |
| Malignancies |  |  |  | 8 |
| Lung | 1 | 1 |  |  |
| Liver |  | 1 | 1 |  |
| Kidney |  |  | 1 |  |
| Colon |  | 1 |  |  |
| Ovary |  | 1 |  |  |
| Unknown primary |  | 1 |  |  |
| Infections |  |  |  | 5 |
| Sepsis | 2 | 2 |  |  |
| TB peritonitis |  | 1 |  |  |
| Mental health-related conditions | 2 |  |  | 2 |
| Unknown | 5 | 4 | 4 | 13 |
| Total | 14 | 19 | 6 | 39 |

## Discussion

We documented a sustained immunorecovery and virologic suppression over 5-year follow-up period. A vast majority of OALHIV in our study (93.1%) who started ART when they were severely immunosuppressed following the treatment guideline in the past could achieve immune recovery. Nearly half had CD4 counts ≥ 500 cells/mm^3^ at the median duration on ART of 8.8 years following treatment initiation. This was in line with the results in the meta-analysis that PLHIV who received ART could survive for more than 10 years after AIDS onset (18). The previous Thai study among participants at the median age of 34.2 years with severe immunocompromised reported a rapid increase CD4 count in the first year and continued to increase at slower rate after 4 years (8). Principally, rapid CD4 count increase immediately after ART initiation is from cell redistribution from lymphoid tissue and peripheral memory T-cell proliferation while thymus attributed to CD4 restoration thereafter. Decreased thymic function with age and low thymus reserve in older adults might cause inferior long-term CD4 restoration compared to younger age group.(19) OALHIV in our cohort might be comparable to the South African study, in which the slower increased CD4 counts after ART initiation in PLHIV >50 years with immunosuppression was observed when compared to the younger age group (10). Nevertheless, the mean CD4 ≥ 500 cells/mm^3^ could be achieved at around 48 months of ART and sustained thereafter in their study. The sustained virologic suppression observed in our study was in line with other Thai studies. The OALHIV cohort in southern Thailand reported 97.9% virologic suppression in 307 patients at 7.27 years follow-up after ART initiation.(20) Another study in Bangkok, Thailand demonstrated 97.4% virologic suppression in 340 OALHIV at 18.4 years after ART initiation (21).

Among OALHIV in our cohort, the probability of 5-year OS was 89.2%. According to the meta-analysis, 4- and 6-year survival probabilities of progression from AIDS onset to AIDS-related death in PLHIV received ART were estimated as 86% and 78%, respectively (18). We started following OALHIV when most of them were considered as stable on treatment. Thus, it was not surprising to see higher probability of survival. A significant lower probability of 5-year survival was seen among OALHIV with CD4 counts < 200 cells/mm^3^ at study entry when compared to those with higher CD4 counts. This was similar to the Ugandan study reported that CD4 counts < 100 cells/mm^3^ was associated with high rate of death in OALHIV (22). The previous study in Thailand among PLHIV at the median age of 33 years, and the median follow-up time of 50 months reported probability of survival was 93.5% at 5 years (23). They mentioned that low CD4 was associated with early mortality in the first 6 months of treatment, while HIV RNA > 1000 copies/mL at 6 months after ART initiation was one among factors associated with long term mortality. However, our population was at older age and had much longer duration on treatment. The median age at death was 62 years was much earlier than life expectancy of general population in this country. According to the World Health organization data published in 2018, the life expectancy in Thailand was 75.5 years, with HIV/AIDS as the 11^th^ causes of death (24). The most identifiable causes of death our cohort were not AIDS, but organ failures and malignancies. Prior to ART and during the first decade after availability of ART, causes of death in PLHIV were mostly attributable to AIDS-defining illnesses as reported from the United Kingdom national observational cohort between 1997-2012, 58% of PLHIV died of AIDS-defining illnesses and late diagnosis was a strong predictor of death (1). Later increasing proportion of death due to non-AIDS disorders such as liver disease, cardiovascular diseases, and malignancies were reported. Data from the antiretroviral therapy cohort collaboration (ART-CC) in Europe and North America reported that rate of AIDS death decrease with time since ART initiation, while mortality from non-AIDS malignancy increased (12).

Despite effective ART, we found that OALHIV faced other health challenges included age-related conditions, and non-AIDS illnesses. The excess mortality and morbidity could be preventable and/or manageable if detected and intervened in a timely manner. At present, HIV RNA level is the only conventional biomarker in HIV treatment monitoring. Meanwhile, a favorable HIV RNA only indicate low risk of HIV treatment failure, but not predictive for overall health in ageing population during long-term follow-up. Moreover, increasing evidence supported that people had a vulnerability to adverse outcomes when facing stressors known as frailty, which could be seen in OALHIV earlier than people without HIV. Regular screening to identify those at risk before they become symptomatic might be useful.

The strength of this study is that we described characteristics and mortality of OALHIV who received HIV care in community hospitals, while other studies mostly conducted in research setting or tertiary care facilities. However, there were several limitations that merit to be addressed. First, we have no baseline data of OALHIV prior to ART initiation. Second, we did not have younger PLHIV in the same setting for comparison, and also no age-matched population without HIV to compare the frequency of age-related comorbidities. Third, the cause of death was incomplete as we could not access to medical record or obtain their health status around the time of death in many cases, despite interview with their relatives.

In summary we documented stable HIV treatment outcome in OALHIV on ART who received HIV care under the national program for more than a decade. Most causes of death were not related to HIV when they were virologic suppressed. The finding supports the need for detection of non-communicable age-related conditions which can occur earlier in OALHIV than general population, to identify those at risk for morbidity and mortality from other diseases. Monitoring of organ functions, cancer surveillance, and mental health screening should be incorporated into HIV programs for aging PLHIV.

## Data Availability

All relevant data are within the manuscript and its Supporting Information files.

## Acknowledgment

We would like to thank all older adults and their relatives who devoted time to participate in this study. We also would like to thank the study staff who conducted data collection, and the HIV clinic staff in community hospitals who helped in reviewing medical records, and coordinating study activities while providing routine services.

